# Cytomegalovirus serostatus and plasma MCP-1 levels are associated with antibody response to seasonal influenza vaccine across age and sex

**DOI:** 10.64898/2026.03.15.26348451

**Authors:** Tamar Ratishvili, Iana H. Haralambieva, Krista M. Goergen, Inna G. Ovsyannikova, Harry Pickering, Matteo Pellegrini, Monica Cappelletti, Elaine F. Reed, Gregory A. Poland, Richard B. Kennedy

**Author notes:** **Address correspondence to**: Richard B. Kennedy, Ph.D., Director, Mayo Vaccine Research Group, Mayo Clinic, Guggenheim 611D, 200 First Street SW, Rochester, Minnesota 55905.

## Abstract

**Background:** While immunologic aging impacts immune responses to vaccination, consistent biomarkers associated with aging of the immune system and suboptimal serologic response to influenza vaccination have not been well-studied. Identification of readily measurable biomarkers of immunosenescence may have predictive clinical utility and inform targeted influenza vaccination strategies and future research into aging of the immune system.

**Methods:** We quantified multiple serum/plasma and cell-based parameters related to immune aging (CMV serostatus, plasma cytokines/chemokines, TREC, TERT, NK cell functionality, and DNA methylation clock) at baseline in an adult (age range 18-85) cohort of 2019-2020 influenza vaccine recipients (n=337) and evaluated their associations with vaccine-induced HAI response to influenza A/H1N1, A/H3N2 and B/Victoria strains.

**Results:** CMV IgG titers were significantly positively correlated with vaccine-induced increases in HAI antibody titers to influenza A/H1N1 (*p=*0.02) and A/H3N2 (*p*=0.014). CMV IgG titers (*p=*0.00096) and CMV seropositivity (*p=*0.003) were also associated with Day 28 HAI seropositivity against influenza A/H3N2 in subjects seronegative at baseline. Conversely, plasma MCP-1 levels were negatively associated with HAI responses to the A/H3N2 (*p=*0.04) strain. These findings were significant independent of age, sex or vaccine type received (high vs standard-dose seasonal influenza vaccine)

**Conclusions:** Our identification of significant relationships between easily quantifiable immune markers and HAI responses to influenza A vaccine strains across sex and age enhances our knowledge of specific links between immune aging and influenza vaccine-induced immunity. These markers could be leveraged for predicting response to influenza immunization.

## Introduction

Influenza is estimated to cause 120,000-710,000 hospitalizations and 6,300-52,000 deaths annually in the United States. [1]Adults ≥65 years are at a disproportionately high risk of adverse influenza-related outcomes and account for 50%-70% of hospitalizations and 70%-85% of deaths during an average influenza season. [2]This situation is aggravated by decreased influenza vaccine responsiveness with older age. [3]Disparities in influenza vaccine-induced immune responses and clinical outcomes in older adults persist despite the use of the high-dose (HD) and MF59-adjuvanted influenza vaccines in this population. [4–6]This highlights the need for a deeper understanding of mechanisms linking aging with influenza-specific immunity to inform the design of novel, targeted influenza vaccines overcoming age-related declines in immune function.

Impaired induction of protective immunity following vaccination, reflected in suboptimal hemagglutination inhibition (HAI) responses (the correlate of influenza vaccine-induced protection) and vaccine effectiveness, has been attributed to immunosenescence. [3, 7]Immunosenescence is the collective term for diverse, usually aging-related, adverse perturbations in the immune system leading to impaired responses to infections and vaccinations. [8]Nevertheless, immunosenescence is not synonymous with chronological aging and reflects immunologic aging, which can progress at a different rate than chronological aging. [9]Ideally, immunosenescence studies would identify quantifiable biologic markers predicting the negative effect of immunosenescence on vaccine-induced immune responses and thus serve as new, useful correlates of vaccine immunogenicity. Such markers could be utilized in new targeted vaccine development including adjuvants, vaccine trials, and clinical decision-making.

We and others have shown that immune aging markers such as increased CD28-T cell frequency, decreased T cell receptor excision circle (TREC) counts, telomerase reverse transcriptase (TERT) activity, and chronic cytomegalovirus (CMV) infection are associated with inter-individual variations in influenza vaccine-induced humoral responses in older individuals. [10, 11]Although helpful, the existing evidence is characterized by: relatively limited selection of markers, enrollment of only older individuals in studies, a focus on a single vaccine strain (e.g., H1N1), and conflicting results on the impact of specific markers. Thus, a comprehensive evaluation of the multipronged impact of immunosenescence on influenza-specific immunity across the age spectrum and across different influenza vaccine virus strains is lacking.

To this end, we assessed a broad panel of cellular and serum/plasma parameters thought to be plausible markers associated with immune aging/immunosenescence to evaluate their relationship with vaccine-induced humoral immunity to influenza A (A/H1N1, A/H3N2) and influenza B (B/Victoria) strains included in the seasonal influenza vaccine. Besides plasma cytokines/chemokines and CMV serostatus, we quantified markers encompassing distinct biological dimensions of immune aging, including thymic output of naïve T cells (TRECs) and cellular replicative capacity (TERT expression), which mainly reflect immunosenescent changes in the T cell compartment, potentially affecting humoral responses to influenza vaccination. We also assessed the associations of Horvath’s epigenetic clock (DNA methylation–derived DNAm age) which estimates biological age based on DNA methylation patterns of selected genes[12], and age-associated functional alterations in NK cells with influenza vaccine-induced immune responses. [13]

In a cohort of younger and older adults, we examined these parameters for their value in predicting influenza-specific immunity and serologic protection.

## METHODS

### 2.1 Ethics Statement

Written informed consent was obtained from all study participants, and the study procedures were approved by the Western Institutional Review Board and the Institutional Review Board of the University of Georgia (IRB #3773).

### 2.2 Study Cohort and Design

The current study utilized demographic data, sera, and PBMCs collected from a large human cohort recruited at the University of Georgia Clinical and Translational Research Unit (Athens, GA) during the 2019-2020 influenza season (UGA4), as previously described. [14–16]Participants received a standard (SD; FLUZONE quadrivalent) or high-dose (HD; FLUZONE trivalent) inactivated seasonal influenza vaccine containing A/H1N1 (A/Brisbane/02/2018), A/H3N2 (A/Kansas/14/2017), B/Victoria (B/Colorado/6/2017-like strain), and [17] B/Yamagata (B/Phuket/3073/2013) influenza strains. Blood was collected before (baseline, D0) and 28-35 days after vaccination (D28). We evaluated multiple markers of immunosenescence in plasma/serum, PBMCs, or DNA and RNA extracted from PBMCs. Due to the limited availability of PBMCs, cell-based markers were quantified only in a subset of study participants. Study design, number of data points available per parameter and assays are outlined in **Figure 1a**.Although the term is imprecise due to the lack of definitive, universally accepted biomarkers, for consistency, we use the terms “markers of immune aging” throughout to refer to candidate indicators considered relevant to immunologic aging.

**Figure 1.**
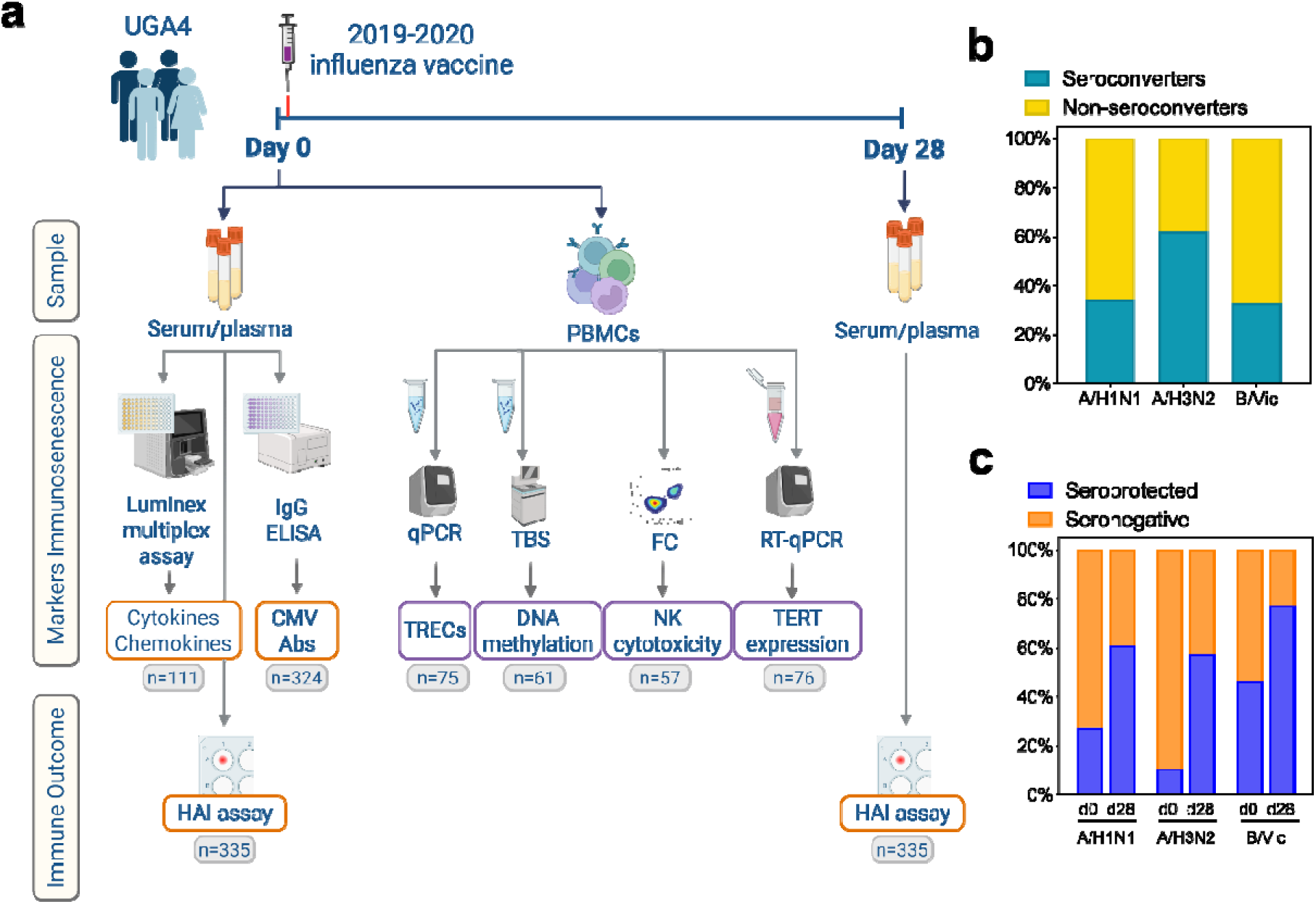
Experimental design and responses to influenza vaccination. **(a)** The cohort of 337 individuals sampled in Athens, GA (UGA4 cohort) received 2019-2020 seasonal influenza vaccination. Blood was collected on Day 0 (before vaccination) and Day 28-35 after vaccination. Influenza-specific humoral immune responses were quantified in serum by HAI assay at both time points. Markers of immunosenescence were assessed in plasma/serum or PBMC samples processed from blood collected at baseline, in varying numbers of subjects as indicated under each outcome. **(b)** The proportions of seroconverting *vs* non-seroconverting subjects to each strain in response to influenza vaccination (D0 to D28); *seroconversion*: ≥4-fold increase in HAI titers and HAI titer ≥1:40 after vaccination. **(c)** The proportions of seroprotected *vs* seronegative subjects to each strain at D0 and D28; *seroprotection*: HAI titer ≥1:40 (at a given time point). CMV, cytomegalovirus; FC, flow cytometry; NK, natural killer; RT-qPCR, reverse transcription - quantitative polymerase chain reaction; TERT, telomerase reverse transcriptase; TRECs, T cell receptor excision circles.

### 2.3 Hemagglutination Inhibition (HAI) Assay

HAI titers specific to each influenza virus strain (A/H1N1, A/H3N2, B/Victoria strains matching the seasonal vaccine composition) were evaluated in subjects’ sera (n=335) at two time points at University of Georgia using previously extensively detailed methods. [14, 18]

### 2.4 Cytomegalovirus (CMV) IgG Antibody EIA

CMV-specific IgG antibodies were assessed in subjects’ sera (n=324) using commercial CMV IgG EIA kits (Bio-Rad Laboratories, Inc., Redmond, WA) following the manufacturer’s instructions (**Supplementary Information *1.1***). Antibody titers are expressed as average index values, with values <0.9 indicating a negative result for CMV IgG antibodies; ≥1.1 are interpreted as positive, while values ≥0.9 and <1.1 are considered equivocal.

### 2.5 Quantification of T cell receptor excision circles (TRECs)

TRECs, non-replicative extrachromosomal DNA remnants of TCR rearrangement, were quantified by real-time qPCR at the Mayo Clinic Cellular and Molecular Immunology Lab using a clinical TREC assay in DNA extracted from PBMCs (n=75) (**Supplementary Information *1.2*)**. Cell count-normalized TREC copy numbers are expressed as copies per million PBMCs.

### 2.6 Assessment of telomerase reverse transcriptase (TERT) expression

TERT RNA levels were measured via qRT-PCR in total RNA extracted from PBMCs (n=76) using Qiagen RT2 SYBR Green/ROX qPCR Mastermix (details provided in **Supplementary Information *1.3***) TERT Ct values normalized to GAPDH levels are represented as a fold-change compared to the reference sample TERT expression using the standard 2^-ΔΔCT^ method.

### 2.7 NK cell cytotoxicity assay

NK cell cytotoxicity was measured by evaluating *in vitro* killing of K562 cells expressing GFP (ATCC® CCL-243-GFP™) co-cultured with subjects’ PBMCs (n=57) using an optimized flow cytometry-based protocol. Details are provided in **Supplementary Information *1.4*.**

### 2.8 Measurement of plasma cytokines and chemokines

Plasma cytokines and chemokines were measured using human 38-plex magnetic cytokine/chemokine kits (EMD Millipore, HCYTMAG-60K-PX38) following previously described methodology (n=111).[19] The Luminex assay and analysis were completed using Luminex 200TM at UCLA Immune Assessment Core. Analyte concentrations (pg/mL) were computed using Milliplex Analyst software version 4.2 (EMD Millipore). Only the analytes detectable (≥ lower limit of detection [LLOD]) in at least 75% of samples were included in the analyses, including GM-CSF, IFNγ, IL-10, IL-1RA, IL-8, MCP-1, and TNFα. For these analytes, values below the LLOD were set to LLOD for downstream analyses.

### 2.9 DNA methylation and Horvath’s clock

DNA methylation was analyzed using targeted bisulfite sequencing (TBS-seq) of DNA extracted from PBMCs of study subjects (n=61) using the phenol-chloroform method at UCLA, as thoroughly described recently.[20] Epigenetic DNA methylation (DNAm) age was calculated using Horvath’s clock[12] using the methylclock R package (**Supplementary Information *1.5***).[21]

### 2.10 Statistical Analysis

Seroprotection is defined as HAI titer ≥ 1:40 and seroconversion as a ≥ 4-fold increase in HAI post-vaccination (Day 0 to Day 28) and a post-vaccination HAI titer ≥ 1:40. To analyze the overall serologic response to seasonal influenza vaccination, we calculated maximum Residual after Baseline Adjustment (maxRBA) using the *titer* R package (https://bitbucket.org/kleinstein/titer). [22] This method accounts for variability in baseline HAI titers and thus preexisting immunity by modeling the expected post-vaccination response as a function of baseline titers and defining the maximal residual across vaccine strains as the individual’s response score. [22–25] The relationships between markers were assessed using Spearman’s rank correlations. Wilcoxon-Rank Sum tests were used to evaluate differences in immunosenescence markers and immune responses between specified groups. Univariate linear regression models were run with maxRBA as the response and markers of immune aging as the predictor variables, adjusting for age, sex, and vaccine dose. For markers significantly correlated with maxRBA score, additional adjusted linear regression models were run for the log2 fold changes for each strain (e.g. Log2(H1N1 Day 28/Day 0), “*HAI response”*). To test the associations of these significant markers on maxRBA by age (young/old) and by sex, linear regression models with interaction terms were used. Similarly, univariate logistic regression models were run using the same predictors and adjustments as above, with HAI seropositivity as the response variable. Analyses were run using R version 4.5.0.

## RESULTS

### Study cohort characteristics and influenza vaccine-induced humoral immune response outcomes

The study design is summarized in **Figure 1a**. Immune response outcomes and other data from subsets of this cohort (UGA4) have been previously reported. [18]Our study subcohort consisted of 337 generally healthy, community-dwelling adult recipients of the 2019-2020 seasonal influenza vaccine, age 18 to 85 years (median age 51 [IQR, 38; 65]; 60.7% female; 88.3% White). Per vaccination recommendations for older individuals, sixty-nine study participants age ≥65 years received the high-dose trivalent Fluzone, which did not include the B/Yamagata strain, and thus B/Yamagata immune outcomes are excluded from all our analyses. As reported previously[18], influenza-specific HAI titers varied in magnitude and by viral strain (**Figure 1b, 1c; Supplementary Table 1**). A large proportion of participants were not seroprotected at baseline (HAI titer <1:40) and potentially susceptible to influenza infection (**Supplementary Table 1**). As expected, vaccination increased the seroprotection rates against each strain on Day 28 (**Figure 1c**). Study cohort demographics and HAI responses are further detailed in **Figure 1b-c**, and **Supplementary Table 1**.

### Markers of immunosenescence and their interrelationships

Measures of immunosenescence assessed in plasma/serum and PBMCs **(Figure 1a)** were widely distributed (**Supplementary Table 2**) and significantly correlated with each other (**Figure 2a**). Plasma MCP-1, TNFα, eotaxin, and IFNγ levels were correlated with age (r=0.39, *p*=2.93×10^-5^; r=0.28, *p*=0.003; r=0.25, *p*=0.008; r=-0.20, *p*=0.03). As expected, TREC content, reflecting thymic output of new T cells, and the proportion of CD56bright NK cells, specifically in the CD16- population, decreased with age (r=-0.55, *p*=4.24×10^-7^ and r=-0.31, *p*=0.018). Conversely, the proportion of total CD56+ NK cells and CD56dim (in both CD16+ and CD16- populations) NK cells increased with age (r=0.27, *p* = 0.04, and r=0.29, *p*=0.03) (**Figure 2a**).

**Figure 2.**
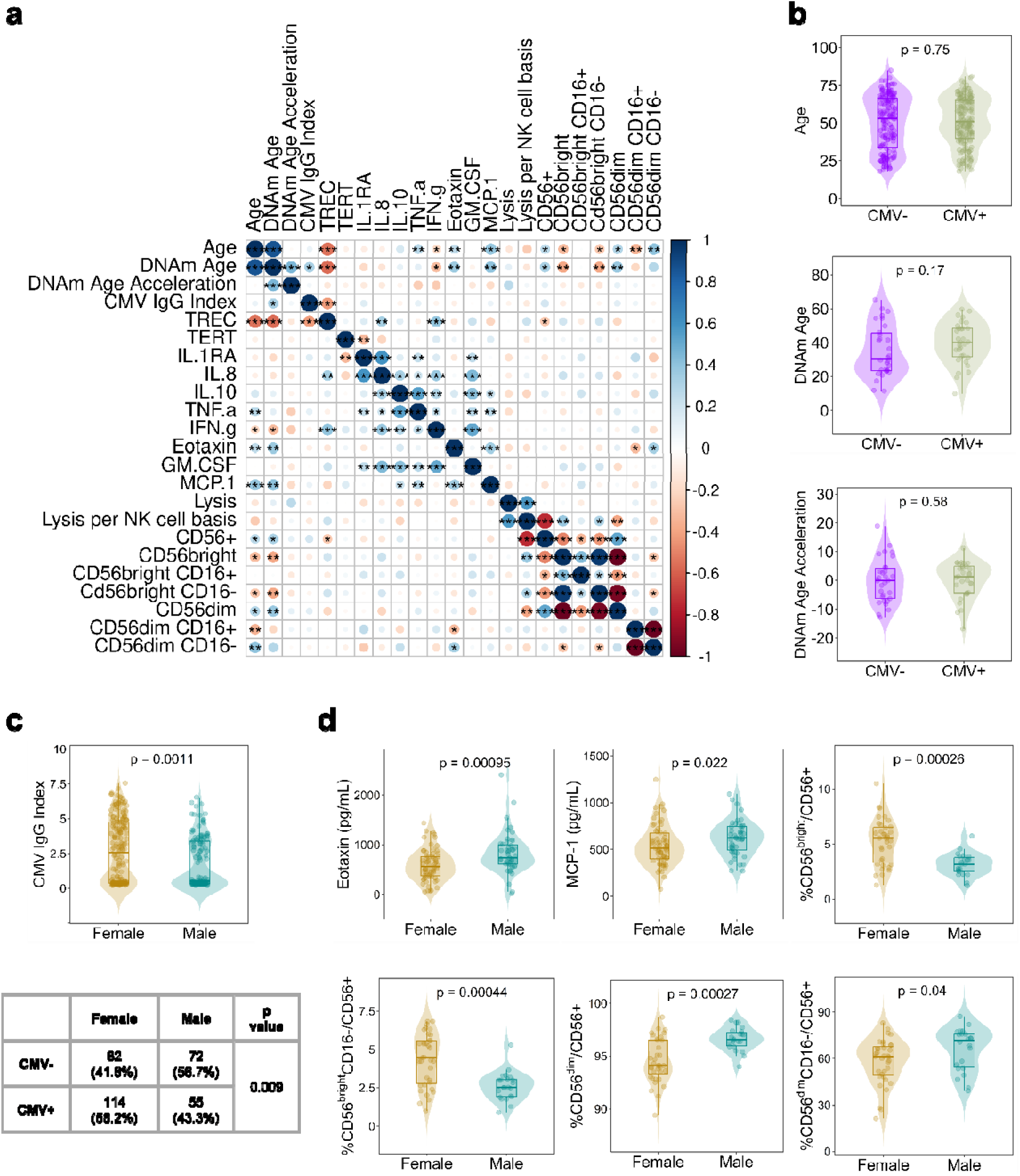
Markers of immunosenescence. **(a)** Correlation matrix between markers of immunosenescence. Color scale and dot size correspond to Spearman’s correlation coefficients (r), with blue shading indicating positive and red shading indicating negative correlation. Asterisks correspond to *p-*values: * <0.05, **<0.01, ***<0.001. **(b)** Comparison of age (top panel), DNAm age (middle panel), and DNAm age acceleration (bottom panel) between CMV seropositive and seronegative individuals. Differences between groups are calculated by Wilcoxon rank-sum test. **(c)** Differences in CMV IgG levels (average sample index values; Wilcoxon rank-sum test; upper panel), and CMV serostatus (CMV+ *vs* CMV-; Chi-square test) between males and females. **(d)** Differences in markers of immunosenescence between males and females. Note: only the markers with significant sex differences besides CMV IgG and seropositivity (panel c) are displayed. *p* values <0.05 are considered statistically significant.

Epigenetic DNAm age was highly correlated with chronological age and consequently, with age-associated markers of immunosenescence (**Figure 2a**). DNAm age was positively correlated with CMV IgG titers (r=0.32, *p*=0.01), and CMV+ individuals tended to be of older DNAm age compared to CMV- individuals (*p*=0.17) (**Figure 2a, 2b)**.

Plasma levels of many cytokines and chemokines were highly correlated (**Figure 2a**) as were NK cell phenotypes, due to their mutually exclusive gating (**Figure 2a**). Individual correlations are detailed in **Supplementary Table 3.**

### Sex-based differences in markers of immunosenescence

We evaluated the impact of biological sex, a variable known to have a multidimensional effect on immunity, on quantified markers of immunosenescence. Females had higher CMV IgG titers (median average index value 2.6 *vs* 0.5, *p*=0.001), and a greater rate of CMV seropositivity (CMV+) (58.2% vs 43.3%, *p*=0.009) (**Figure 2c**). IgG titers were also higher in CMV+ females than in CMV+ males (median 4.5 *vs* 3.5, *p=*0.003). Plasma Eotaxin (median 734.3 *vs* 564.4 pg/ml, *p*=0.0009) and MCP-1 levels (median 621.1 *vs* 512.7 pg/ml, *p*=0.02) were higher in males than females. Women also had higher frequencies of *CD56bright* NK cells (median 5.5% *vs* 3.2%/live CD56+ cells, *p*=0.0003) (**Figure 2d**). The distributions of all assessed markers between sexes are detailed in **Supplementary Table 2**.

### Relationship between demographic factors and humoral immune response outcomes following influenza vaccination

Our initial analysis used the maxRBA score to capture the relationship between independent variables and overall influenza vaccine-induced immune response adjusted for inverse correlation between pre-existing HAI titers and post-vaccination HAI fold-change. [22]This was followed by a secondary analysis to assess the relationships with strain-specific vaccine-induced HAI response (log2[D28/D0]) (see *Statistical Analysis*).

To address differences in the vaccine formulation received (SD vs HD) by age, additional analyses were performed to assess the independent effects of age and vaccine formulation on HAI responses. In univariate linear models in the whole cohort, both age and HD vaccine formulation were significantly negatively associated with maxRBA (**Supplementary Table 4, Supplementary Figure 1a**). However, in a multivariable model including age, sex and vaccine dose, only age remained significantly, albeit modestly, associated with maxRBA (**Figure 3a, Supplementary Table 4**). In a subgroup analysis of adults ≥65 years, the only group receiving both SD (n=21) and HD (n=69) vaccines, maxRBA and per-strain post-vaccination log□ HAI responses were comparable between recipients of different vaccine formulations (**Supplementary Figure 1b**, **Supplementary Table 5**). Further, baseline characteristics, such as age, sex and BMI, did not differ between the two groups of older adults (**Supplementary Table 5**). Similarly, among SD recipients in the whole cohort, immune response outcomes did not differ between adults <65 and ≥65 years of age (**Supplementary Figure 1c**). Overall, as these findings suggest that age, rather than vaccine formulation accounts for variations in post-vaccination influenza-specific responses, both age and vaccine recipient groups were analyzed together hereafter.

**Figure 3.**
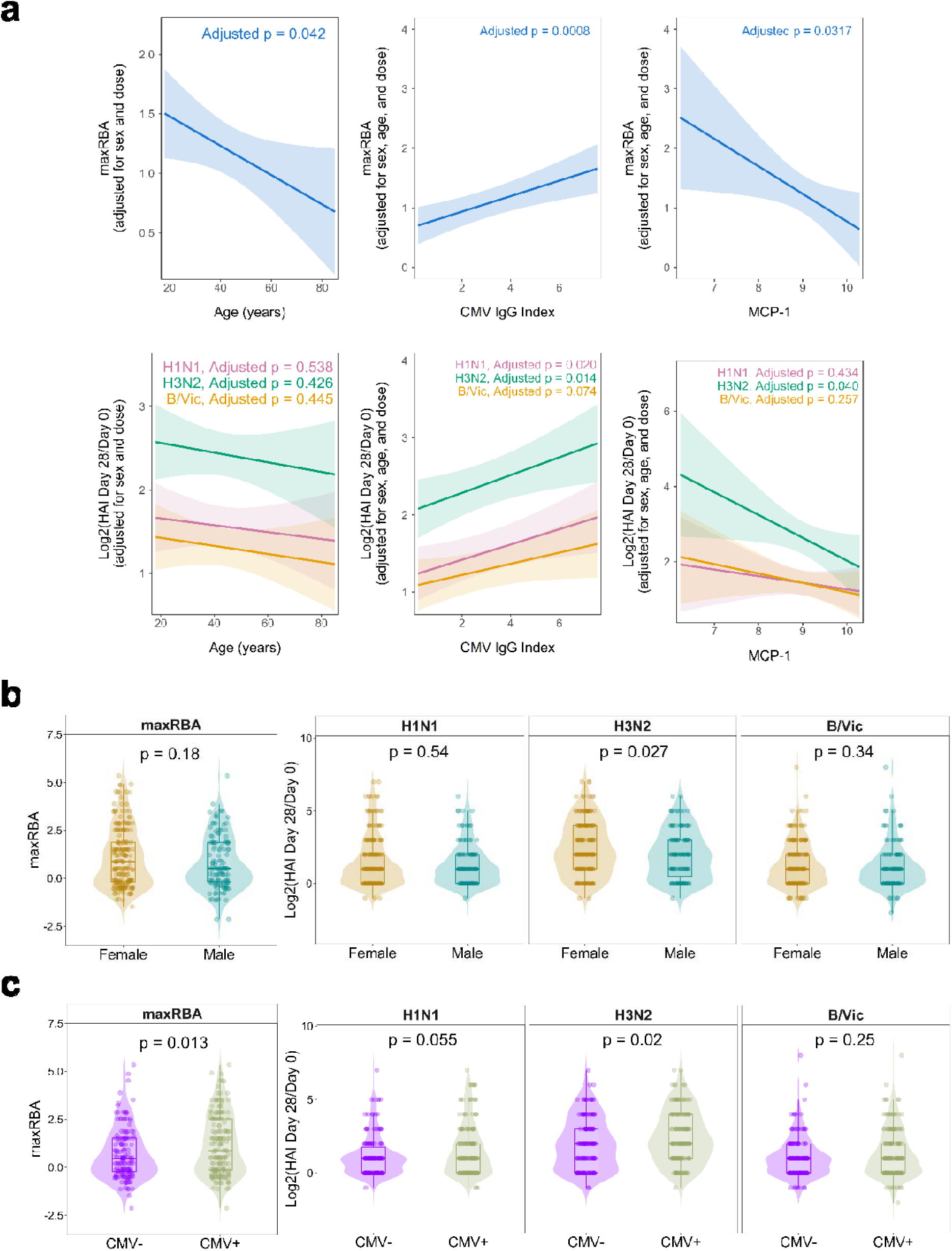
Markers of immunosenescence and HAI responses. **(a)** Linear models predicting maxRBA or log2 fold-change in HAI titers to each influenza strain following vaccination (D28-D0) adjusted for received vaccine dose, sex, and age. Each line corresponds to each strain, shading indicates 95% confidence intervals. **(b)(c)** Differences in maxRBA (left panels) and log2 fold-change in HAI titers to each strain following vaccination (D28-D0) (right panels) by sex **(b)** or CMV serostatus **(c)**. Solid lines correspond to medians, and boxplots span the interquartile range. *P* values are calculated using the Wilcoxon rank-sum test. *p* values <0.05 are considered statistically significant.

Further, per-strain HAI responses were not significantly associated with age (**Figure 3a**), while females demonstrated a greater HAI response to A/H3N2 (*p=*0.027) (**Figure 3b**) but not A/H1N1 or B/Victoria.

### Associations between markers of immune aging and humoral immune response outcomes following influenza vaccination

Unexpectedly, we found a significant direct association between CMV IgG levels and maxRBA (*p*=0.00075) (**Figure 3a**), HAI response to influenza A/H1N1 (*p*=0.02) and A/H3N2 (*p*=0.014), and a marginal positive correlation with B/Victoria (*p=*0.07) (**Figure 3a**). Similarly, CMV+ individuals had significantly higher maxRBA scores (*p*=0.013), and HAI responses to A/H3N2 (*p*=0.02) and marginally to A/H1N1 (*p*=0.055) than CMV- participants (**Figure 3c**). Further, CMV IgG titers were associated with vaccine-induced seroprotection (D28 HAI titer ≥ 1:40 among individuals seronegative at D0) against A/H3N2 (*p*=0.00096) when adjusted for age, sex, and vaccine dose (**Supplementary Table 6**), with CMV+ individuals more likely to attain seroprotection than CMV- individuals (*p*=0.003, **Supplementary Table 7**).

Conversely, baseline plasma MCP-1 levels were negatively associated with maxRBA score (*p*=0.032), and HAI response to A/H3N2 (*p*=0.04) (**Figure 3a**).

As CMV serostatus was associated with sex and plasma MCP-1 levels correlated with both sex and age, we included the interaction terms of age ≥65 years and male sex in models predicting maxRBA to evaluate potential differences by sex and age group. No significant interactions between MCP-1 and age groups or sex were identified **(Supplementary Table 4)**, indicating that the associations with MCP-1 were consistent across sexes and age groups. Similarly, no significant interactions were observed between CMV IgG and age group, although the interaction between CMV IgG and male sex was marginally significant (p = 0.056) **(Supplementary Table 4)**, suggesting a stronger association between CMV IgG levels and maxRBA in males independent of age group. Model outputs are detailed in **Supplementary Table 4**.

## DISCUSSION

In this report, we evaluated a diverse set of markers associated with immune aging. Besides the modest associations of age and sex with the overall HAI response (maxRBA) to influenza vaccination and HAI response to the A/H3N2 strain, respectively, two parameters - CMV IgG antibody titer/CMV seropositivity, and pre-vaccination MCP-1 levels - correlated with vaccine-induced immune responses to influenza in this study (CMV: A/H1N1, A/H3N2; MCP-1: A/H3N2). Even though CMV serostatus and plasma MCP-1 levels were differentially distributed by sex, and MCP-1 levels increased with age, we demonstrated that age did not interact with these markers to predict HAI response to influenza vaccination. However, the relationship with CMV IgG was marginally stronger in males. Thus, our findings are generalizable to individuals independent of their age, suggesting that immunologic aging is a distinct phenomenon beyond chronological aging.

Our finding of a moderate but consistent positive relationship between CMV serostatus/IgG titers and HAI response to influenza A vaccine strains seemed counterintuitive due to the assumed negative role of CMV in immune aging. While latent CMV infection clearly relates to one of the immune aging hallmarks, i.e., shift of T cell subset distribution away from naïve cells and towards late stage, highly differentiated memory phenotypes with decreased expression of CD28/upregulation of CD57, evidence suggests that CMV serostatus does not universally have an *in toto* negative influence on immune responses to vaccination. [26–28]In fact, a comprehensive meta-analysis challenges the assumption of a negative influence of latent CMV on influenza vaccine-induced humoral immune responses, demonstrating that existing data are inconsistent or conflicting. [26]Specifically, several studies demonstrated negative correlations between CMV seropositivity and humoral immune response outcomes (mostly HAI titers) to influenza vaccination. [29–33]Some of them linked this relationship to the increases in terminally differentiated CMV-specific T cells (which we did not measure) [30, 32, 34]and/or demonstrated this relationship only in young [29] or older vaccinees. [32, 33]In contrast, other reports, including our current study, demonstrate a positive correlation between CMV seropositivity and influenza vaccine-induced HAI response depending on time since vaccination[27, 28, 35], while yet others fail to establish any association. [28, 36, 37] Positive relationships were more often shown in young individuals[26], but our study did not demonstrate an age-dependent association. These divergent findings indicate that other factors pertinent to study design and/or host response to CMV and/or to influenza, such as preexisting immunity/original antigenic sin, potential differences in CMV’s impact on recall *vs de novo* antigen response, and, clearly, T cell-mediated immune response, likely modulate this relationship. Nevertheless, a study utilizing the then-newly emerged H1N1pdm09 strain to address this issue did not identify a negative impact of CMV on *de novo* vaccine-induced HAI responses after 3 and 6 weeks following vaccination, while CMV+ individuals demonstrated significantly higher HAI responses at 26 and 52 weeks. [28]

Information on the impact of CMV on influenza-specific B cells and immunosenescence is scarce. A study examining the B cell repertoire revealed higher fractions of long-lived B cell lineages responding to influenza vaccination (B cell heavy-chain transcript increase from Day 0 to Day 7 post-vaccination) in old CMV+ compared to young CMV- subjects, concluding that latent CMV infection is associated with greater activation of existing antigen-specific memory B cells in both young and old. [38]Further, host immune response to CMV including pro-/anti-inflammatory cytokine and Th1/Th2 bias,[39] viral load, reactivation history, and time since first exposure, likely also modulate the effect of CMV on the immune response. This suggests that CMV serostatus/IgG titers quantitated at a single point in time, do not fully capture CMV burden on immunity. [40] Moreover, CMV serostatus and IgG titers differed between sexes in our study, and a previous report demonstrated sex-based differences in CMV-induced redistribution of T cell subsets with CMV+ males demonstrating lower numbers of memory CD4 Th and B cells[41]. Our models suggested marginally male-biased positive effect of CMV on HAI response in the UGA4 cohort, warranting further investigation. A thorough evaluation of both humoral and cellular responses to CMV, detailed characterization of T cell subpopulations in CMV- and CMV+ females and males, in conjunction with influenza-specific humoral and T cell immune responses to different strains at multiple timepoints after vaccination is needed to uncover the dynamics and mechanisms of the effect of CMV on pathogen-specific adaptive immune responses.

Our study also demonstrated a moderate inverse relationship between baseline plasma MCP-1 levels and HAI responses. MCP-1, monocyte chemoattractant protein 1 or CCL2, is a pro-inflammatory mediator primarily acting through the chemotaxis of monocytes, T cells, NK cells, and DCs in the pathogenesis of several inflammatory and age-related human diseases such as diabetes mellitus, atherosclerosis, and COVID-19. [42] Circulating MCP-1 reflects the combined secretion from monocyte-derived, endothelial, tissue-resident, smooth muscle, and other cell types. [42] Although systemic MCP-1 levels increase with chronological age [17, 43], also observed in our cohort, the negative association of plasma MCP-1 with influenza vaccine-induced response in this study appeared independent of age and sex. This conforms with existing evidence: MCP-1 is not merely a correlate of chronological age, but rather a proposed marker of biological aging. To this point, MCP-1 is expressed at higher levels in sera of frail humans compared to non-frail, age-matched controls and in progeroid mouse models, with the latter responding by reduction in MCP-1 to anti-aging interventions (senolytic treatment). [43]MCP-1 is a prominent component of SASP, or the senescence-associated secretory phenotype [44], a hallmark of senescent cells that likely contribute to age-related immune dysregulation and the chronic low-grade inflammation of aging (“inflammaging”).[45] Chronic inflammatory signaling could impair antigen presentation and T/B cell responses through their effect on bystander cell activation, survival and functionality. [46–48] Thus, elevated baseline MCP-1 may reflect increased systemic aging or inflammatory burden that could impair the ability to mount effective influenza vaccine–induced immune responses.

Several markers examined in our study, including TRECs, TERT expression, NK cell cytotoxicity, and DNAm age and acceleration, were not significantly associated with influenza vaccine-induced humoral immune responses. In our previous study, TRECs and TERT activity were positively correlated with Day 3 and Day 28 post-vaccination A/H1N1-specific memory B cell ELISPOT responses, respectively, but not with HAI responses in adults ≥55 years. [11] TRECs and TERT are indicators of thymic output and lymphocyte proliferative capacity, respectively, which may influence early antigen-specific B-cell activation and affect memory B cell ELISPOT responses but may not always directly translate into circulating HAI titers. Regarding our negative findings with DNA methylation, Horvath’s DNAm age and age acceleration were not different among responders and non-responders to trivalent influenza vaccine in one small study[49]. Future studies should evaluate the utility of other epigenetic clocks[50] as markers of reduced influenza vaccine responsiveness. To the best to our knowledge, *baseline* NK cell cytotoxicity as a marker of immunosenescence has not been previously evaluated against influenza-specific humoral immune responses. It should be noted, however, that these markers were quantified in a smaller sample of study subjects due to limited PBMC availability (n=57-75), and larger studies are needed for drawing firm conclusions.

The strengths of our study include the usage of a wide panel of potential markers of immune aging in a relatively large cohort. Our study also has limitations, including the high heterogeneity/variation in immune response, indicating that validation and replication of our findings in a larger cohort is necessary. Functional studies will also be needed to elucidate biologic mechanisms underlying our identified associations. Further, we focused only on humoral immune responses, and while HAI titers are the gold standard, our findings need to be contextualized with T cell-mediated influenza-specific immunity and immunosenescence-associated changes in T cell subpopulations. This is a subject of our ongoing complementary studies in a separate cohort of older influenza vaccine recipients. Similarly, future studies should include frailty and age-associated comorbidities as covariates for comprehensive insights.

Existing knowledge gaps limit practical utility of markers associated with immune aging in influenza vaccinology research and clinical practice. Our findings suggests that this represents a potential missed opportunity and that these parameters could be used to better predict post-vaccination immunity and devise vaccination strategies to overcome their adverse effects. We conclude that with extended evaluation in larger human cohorts, plasma MCP-1 could potentially be used as an easily quantifiable analyte predicting diminished HAI response to one or more influenza vaccine strains. Our findings provide additional information to the debate on the prognostic value of CMV IgG serostatus in vaccine-induced humoral immune response to influenza A strains, delineating the need for further in-depth studies into the impact of chronic CMV infection on immunity. While it is early to define influenza vaccination-specific biomarkers of immunosenescence that could be translated into clinical trial settings, studies like ours add significant evidence and provide a framework for future functional studies to investigate the biology underlying these associations. These studies could examine the use of adjuvants to overcome immune aging, and ascertain whether these findings are specific to influenza or if they predict vaccine responses more broadly.

## Acknowledgements

We thank Dr. Ted Ross and Michael Carlock (UGA/Cleveland Clinic) for generously sharing biospecimens and HAI data from their study cohort; Dr. Lin Wang, Jack M. Monroe, Ilya M. Swanson and Kyle Calley for performing the experiments. We thank NIAID and CIVICs for funding.

The authors acknowledge the UGA clinical teams for sample collections blood and saliva processing and technical assistance. We would also like to thank all participants enrolled in the study, as well as Dr. Brad Phillips, Kimberly Schmitz, and the entire staff at the University of Georgia Clinical and Translational Research Unit (CTRU) for assistance in collecting samples in the influenza vaccine program. The CTRU was supported by the National Center for Advancing Translational Sciences of the National Institutes of Health under Award Number UL1TR002378.

## Data availability statement

The data presented in this manuscript are included as figures, tables and supplementary materials, and will be made available upon a reasonable request from the corresponding author.

