## Supplementary figures and images for "Cytomegalovirus serostatus and plasma MCP-1 levels are associated with antibody response to seasonal influenza vaccine across age and sex"

### Supplementary Figure 1

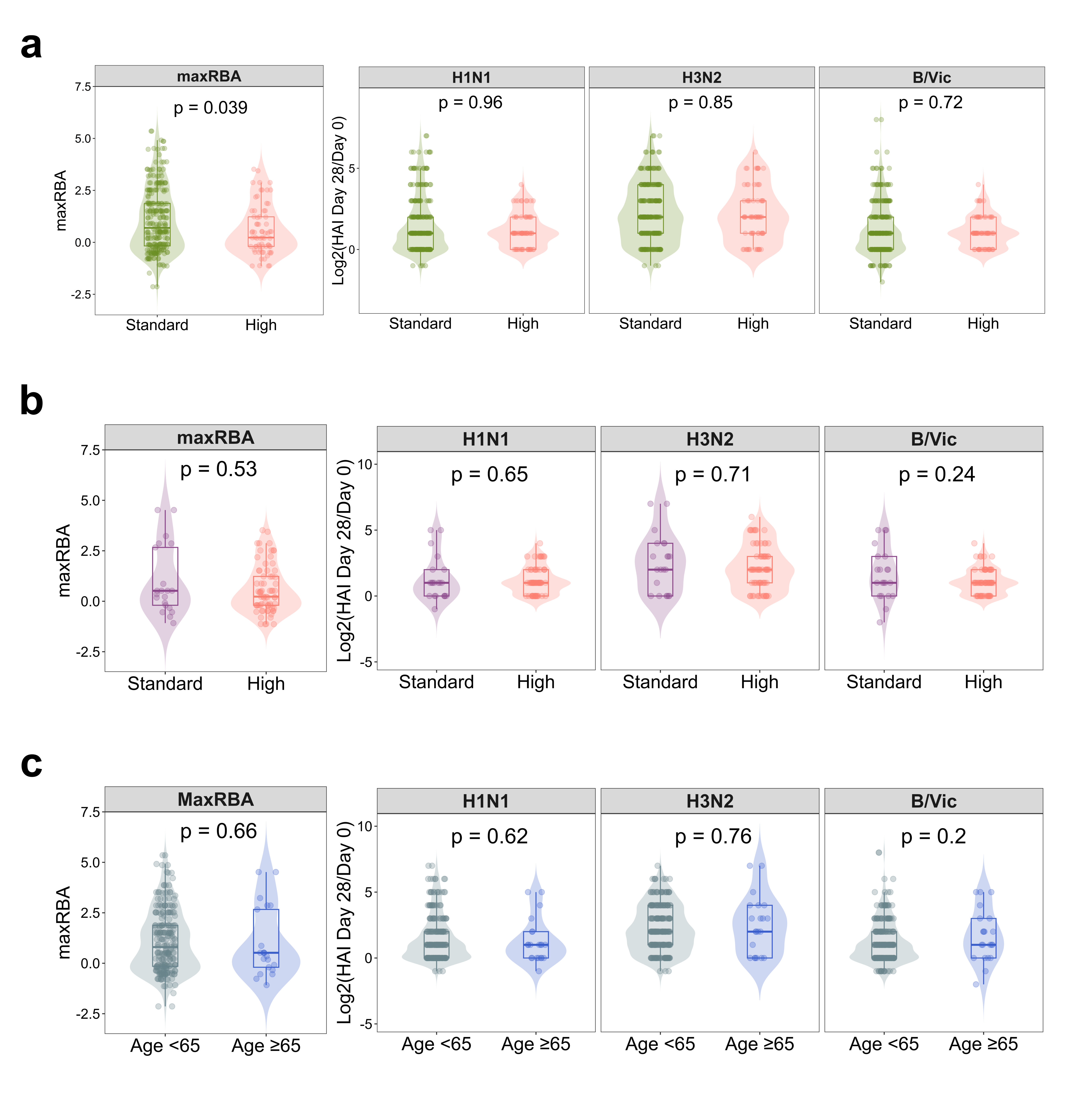
