## Supplementary Figure 1 caption for "Cytomegalovirus serostatus and plasma MCP-1 levels are associated with antibody response to seasonal influenza vaccine across age and sex"

**Supplementary Figure 1.** Comparison of responses to influenza vaccination by received vaccine formulation (Standard vs High Dose). Violin plots depict comparisons in maxRBA and per-strain post-vaccination HAI responses among **a)** all study participants by received vaccine dose; **b)** adults ≥65 years (the only age group receiving HD vaccine) by received vaccine dose; **c)** SD vaccine recipients by age group (<65 years and ≥65 years). Solid lines represent medians; boxplots span the interquartile range (25^th^-75^th^ percentiles; Q1-Q3).

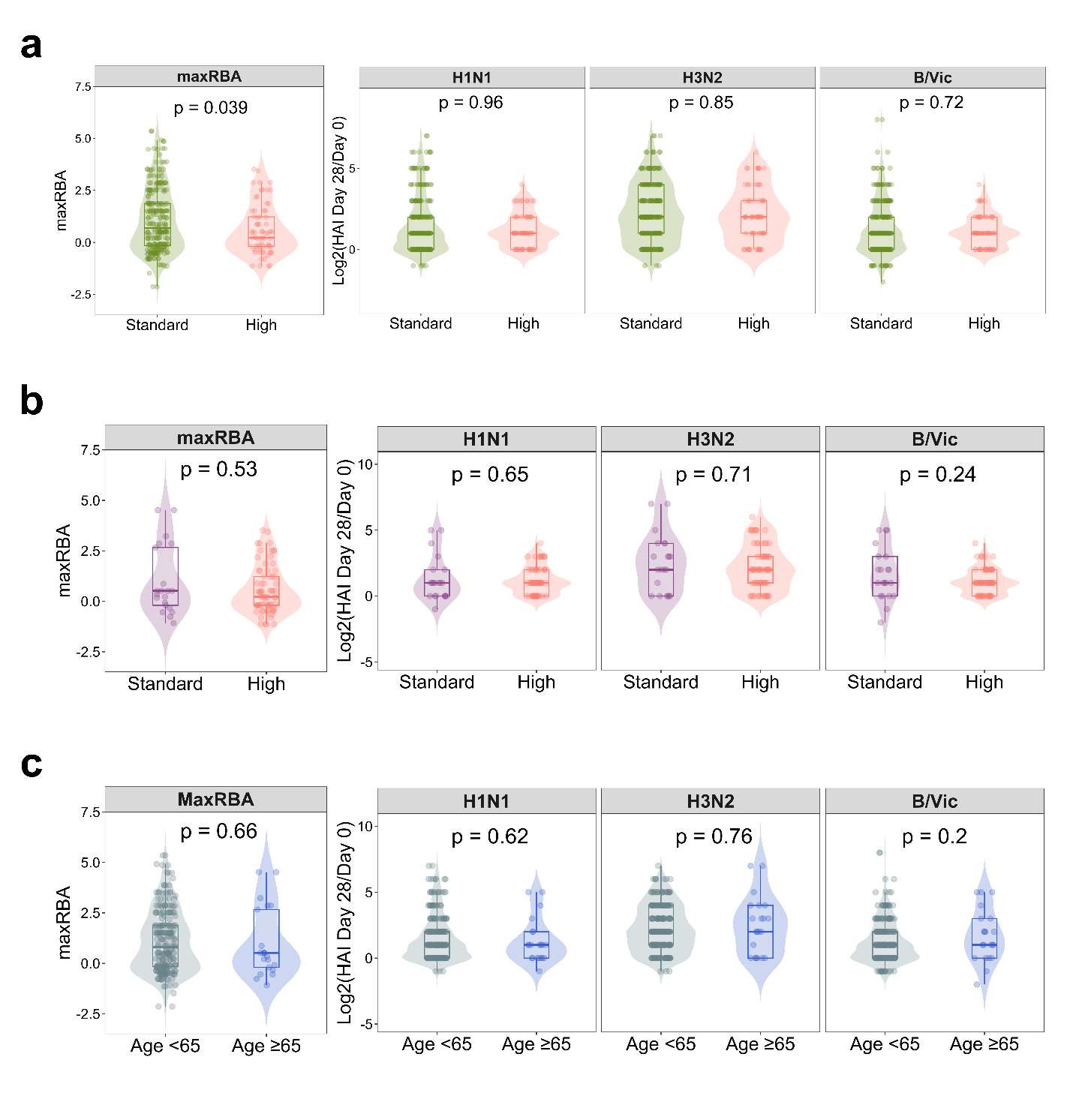
