## Supplementary Methods for "Cytomegalovirus serostatus and plasma MCP-1 levels are associated with antibody response to seasonal influenza vaccine across age and sex"

**Supplementary Information**

1.1 ***CMV IgG Antibody EIA brief description***

Participants’ serum samples and assay controls were added to plates precoated with CMV antigen (strain AD 169). After the incubation, plates were washed, incubated with secondary detection reagents, and optical density was measured. CMV IgG antibody levels are expressed as average index values, with values <0.9 indicating a negative result for CMV IgG antibodies; ≥1.1 are interpreted as positive, while values ≥0.9 and <1.1 are considered equivocal.

1.2 ***Details of TREC assay***

DNA was extracted from thawed cryopreserved PBMCs using the Qiagen AllPrep DNA/RNA Mini Kit per the manufacturer’s recommendations. Genomic DNA and TRECs were quantified in triplicate by real-time PCR using fluorescent probes specific for TCR delta-deletion TREC signal joint and albumin. Absolute TREC and albumin quantities were determined from fluorescence intensity measures using a standard curve. TREC copy numbers were normalized to cell counts determined using albumin counts and expressed as copies per million PBMCs.

1.3 ***Details of TERT quantification assay***

Total RNA was extracted from PBMCs using the Qiagen RNeasy Plus mini kits according to the manufacturer’s recommendations. cDNA was generated via random-primer reverse transcription using the Qiagen RT2 First Strand kit according to the manufacturer’s instructions. qPCR was performed using the ready-to-use Qiagen RT2 SYBR Green/ROX qPCR Mastermix. Quantitative PCR was performed on a Thermo Fisher Scientific QuantStudioTM 7 Pro Real-Time PCR system. The Agilent Quantitative PCR Human Reference Total RNA (Agilent cat. # 750500) was used as a reference.

*Primer sequences*

TERT: OriGene Telomerase reverse transcriptase (TERT) Human qPCR Primer Pair (cat. # HP230441). Forward primer: GCCGATTGTGAACATGGACTACG. Reverse primer: GCTCGTAGTTGAGCACGCTGAA).
GAPDH: OriGene GAPDH Human qPCR Primer Pair (cat. # HP205798). Forward primer: GTCTCCTCTGACTTCAACAGCG. Reverse primer: ACCACCCTGTTGCTGTAGCCAA).

1.4 ***NK cell cytotoxicity assay***

K562-GFP cells were co-cultured with study participants’ PBMCs for 3 hours and stained on ice with a fluorochrome-conjugated antibody cocktail (APC-CD3, BV711-CD16, and BV421-CD56 [BioLegend, San Diego, CA]) for 20 min and centrifuged at 1500rpm for 5 min. Plates were resuspended in viability staining mix (7-AAD in PBS) and acquired at ZE5 flow cytometer (Bio-Rad, Hercules, CA). Data were analyzed using FlowJo software. NK cells were gated based on CD56/CD16 expression on CD3 negative singletons. Lysis (%) of target K-562-GFP cells, reflecting NK cell cytotoxic activity, is represented as a frequency of dead targets (7-AAD+GFP+) among total target cells (7-AAD+/-GFP+). Percent (%) lysis was normalized to each subject’s NK cell frequencies: %lysis / %GFP-CD3-CD56+. [1]

***1.5 DNAm age calculation from targeted bisulfite sequencing data***

DNAm was calculated for the targeted bisulfite sequencing samples by using the UCSC genome browser [https://genome.ucsc.edu/cgi-bin/hgLiftOver] to convert the genome coordinates from NCBI36/hg18 to NCBI38/hg19 for each of the CG sites in the Horvath clock model. For each of the 353 CG sites used in the model, we obtained the percent methylation for the site by finding values within 15 base pairs of the site and then summed over the MC (5-methylcytosine) and NC (non-methylated cytosines) fields and used the MC/NC value.
