## Supplementary Table 1, Supplementary Table 2, Supplementary Table 4, Supplementary Table 5, Supplementary Table 6, Supplementary Table 7 for "Cytomegalovirus serostatus and plasma MCP-1 levels are associated with antibody response to seasonal influenza vaccine across age and sex"

**Supplementary Table 1. Demographic characteristics and HAI antibody titer distributions.** Seropositivity to each strain is defined as HAI titer ≥1:40 at a given timepoint. N-Miss, number of subjects with missing data; Q1, 25^th^ quartile; Q3, 75^th^ quartile; BMI, body mass index.

| Demographic characteristics | | | |
| --- | --- | --- | --- |
|  | Whole cohort (n=337) | By age groups | |
| Age |  | < 65 years (n=247) | **≥** 65 years  (n=90) |
| Median (Q1, Q3) | 51 (38, 65) | 43 (31, 54) | 69.5 (67, 72) |
| Range | 18 - 85 | 18 - 64 | 65 - 85 |
| Sex |  |  |  |
| N-Miss | 1 |  |  |
| F | 204 (60.7%) | 152 (61.8%) | 52 (57.8%) |
| M | 132 (39.3%) | 94 (38.2%) | 38 (42.2%) |
| Race |  |  |  |
| N-Miss | 11 |  |  |
| American Indian/Alaska Native | 2 (0.6%) | 1 (0.4%) | 1 (1.1%) |
| Asian | 7 (2.1%) | 6 (2.5%) | 1 (1.1%) |
| Black | 29 (8.9%) | 26 (11.0%) | 3 (3.3%) |
| White | 288 (88.3%) | 203 (86.0%) | 85 (94.4%) |
| BMI |  |  |  |
| N-Miss | 1 |  |  |
| Median (Q1, Q3) | 27.985 (24.795, 32.15) | 27.992 (24.779, 32.342) | 27.853 (25.183, 31.359) |
| Range | 17.427 - 59.88 | 17.427 – 59.88 | 19.082 – 52.14 |
| Dose |  |  |  |
| N-Miss | 2 |  |  |
| Standard | 266 (79.4%) | 245 (100.0%) | 21 (23.3%) |
| High | 69 (20.6%) | 0 (0.0%) | 69 (76.7%) |

| **Distribution of HAI responses and serostatus** | | | | | | | |
| --- | --- | --- | --- | --- | --- | --- | --- |
| **Outcome** | **Visit** | **N** | **Median** | **IQR** | **Range** | **Seropositive** | **Seronegative** |
| **MaxRBA** | Day 28 - Day 0 | 335 | 0.52 | -0.2, 1.88 | -2.14, 5.35 |  |  |
| **HAI H1N1** | Day 0 | 335 | 10 | 5, 40 | 5, 640 | 91 (27.2%) | 244 (72.8%) |
| **HAI H1N1** | Day 28 | 335 | 40 | 20, 80 | 5, 1280 | 204 (60.9%) | 131 (39.1%) |
| **HAI H1N1** | Day 28 - Day 0 | 335 | 1 | 0, 2 | -1, 7 |  |  |
| **HAI H3N2** | Day 0 | 335 | 5 | 5, 10 | 5, 160 | 35 (10.4%) | 300 (89.6%) |
| **HAI H3N2** | Day 28 | 335 | 40 | 20, 80 | 5, 1280 | 192 (57.3%) | 143 (42.7%) |
| **HAI H3N2** | Day 28 - Day 0 | 335 | 2 | 1, 4 | -1, 7 |  |  |
| **HAI B/Vic** | Day 0 | 335 | 20 | 10, 80 | 5, 640 | 155 (46.3%) | 180 (53.7%) |
| **HAI B/Vic** | Day 28 | 335 | 80 | 40, 160 | 5, 1280 | 259 (77.3%) | 76 (22.7%) |
| **HAI B/Vic** | Day 28 - Day 0 | 335 | 1 | 0, 2 | -2, 8 |  |  |

**Supplementary Table 2. Distributions and sex differences of markers of immune aging.**

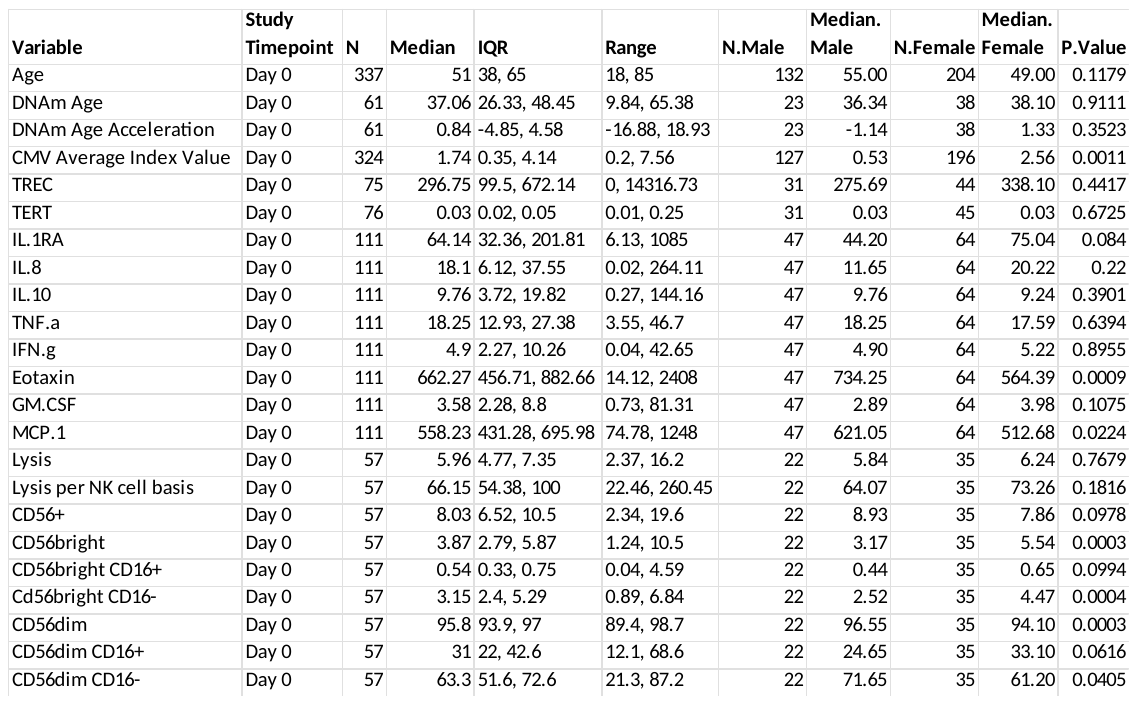

**Supplementary Table 3. Correlations between markers of immunosenescence.** (separate file provided).

**Supplementary Table 4. Linear modeling of demographic factors, markers of immune aging and HAI responses**

| **MaxRBA score by age, vaccine dose and sex model output** | | | | | | | |
| --- | --- | --- | --- | --- | --- | --- | --- |
|  | | | **Unadjusted models** | | | **Adjusted model  (Sex + Age + Dose)** | |
| **Response outcome** | **Predictor** | **Term** | **N** | **Unadjusted β (95% CI)** | **p** | **Adjusted β (95% CI)** | **p** |
| MaxRBA | Age | Age | 332 | -0.015  (-0.025, -0.006) | 0.00135 | -0.012  (-0.024, -0.000) | 0.042 |
|  | Vaccine dose | HD vs SD | 332 | -0.468  (-0.857, -0.079) | 0.0185 | -0.158  (-0.645, 0.330) | 0.526 |
|  | Sex | Male vs Female | 332 | -0.305  (-0.629, 0.019) | 0.0649 | -0.266  (-0.589, 0.057) | 0.106 |

| MaxRBA score by markers, adjusted for sex, age, vaccine dose | | | | | | |
| --- | --- | --- | --- | --- | --- | --- |
| Response  outcome | **Predictor** | **Adjusted** | **N** | **Estimate** | **Std.Error** | **P.Value** |
| maxRBA | **CMV IgG Index** | **Sex, Age, Dose** | **320** | **0.130** | **0.038** | **0.00075** |
| maxRBA | Log2(TREC + 1) | Sex, Age, Dose | 73 | -0.073 | 0.071 | 0.31000 |
| maxRBA | Log2(TERT) | Sex, Age, Dose | 74 | -0.239 | 0.129 | 0.06900 |
| maxRBA | Log2(IL.1RA) | Sex, Age, Dose | 110 | 0.017 | 0.065 | 0.79000 |
| maxRBA | Log2(IL.8) | Sex, Age, Dose | 110 | 0.007 | 0.056 | 0.90000 |
| maxRBA | Log2(IL.10) | Sex, Age, Dose | 110 | -0.100 | 0.066 | 0.13000 |
| maxRBA | Log2(TNFα) | Sex, Age, Dose | 110 | 0.033 | 0.168 | 0.84000 |
| maxRBA | Log2(IFNγ) | Sex, Age, Dose | 110 | -0.020 | 0.054 | 0.71000 |
| maxRBA | Log2(Eotaxin) | Sex, Age, Dose | 110 | -0.121 | 0.113 | 0.28000 |
| maxRBA | Log2(GM.CSF) | Sex, Age, Dose | 110 | -0.005 | 0.071 | 0.95000 |
| maxRBA | **Log2(MCP.1)** | **Sex, Age, Dose** | **110** | **-0.462** | **0.212** | **0.03200** |
| maxRBA | Lysis | Sex, Age, Dose | 55 | -0.017 | 0.079 | 0.83000 |
| maxRBA | Lysis per NK cell basis | Sex, Age, Dose | 55 | -0.001 | 0.004 | 0.90000 |
| maxRBA | CD56+ | Sex, Age, Dose | 55 | 0.017 | 0.050 | 0.74000 |
| maxRBA | CD56bright | Sex, Age, Dose | 55 | 0.006 | 0.106 | 0.96000 |
| maxRBA | CD56bright CD16+ | Sex, Age, Dose | 55 | -0.024 | 0.233 | 0.92000 |
| maxRBA | Cd56bright CD16- | Sex, Age, Dose | 55 | 0.002 | 0.125 | 0.99000 |
| maxRBA | CD56dim | Sex, Age, Dose | 55 | 0.015 | 0.105 | 0.89000 |
| maxRBA | CD56dim CD16+ | Sex, Age, Dose | 55 | 0.004 | 0.014 | 0.79000 |
| maxRBA | CD56dim CD16- | Sex, Age, Dose | 55 | -0.006 | 0.014 | 0.68000 |
| Interaction models of CMV IgG and MCP-1 with sex and age predicting maxRBA | | | | | | |
|  | **estimate** | **std.error** | **p.value** | **adj.r.squared** | | |
| (Intercept) | 0.755 | 0.141 | < 0.001 | 0.039 | | |
| CMV IgG  0.114 | | 0.044 | 0.01 |  | | |
| Age >=65  -0.436 | | 0.259 | 0.094 |  |  |  |
| CMV:Age >=65  0.053 | | 0.083 | 0.525 |  |  |  |
|  | estimate | std.error | p.value | adj.r.squared | | |
| (Intercept) | 0.875 | 0.158 | < 0.001 | 0.045 | | |
| CMV IgG  0.073 | | 0.045 | 0.106 |  | | |
| Sex, Male  -0.579 | | 0.241 | 0.017 |  |  |  |
| CMV:Sex, Male  0.161 | | 0.084 | 0.056 |  |  |  |
|  | estimate | std.error | p.value | adj.r.squared | | |
| (Intercept) | 4.284 | 2.043 | 0.038 | 0.009 | | |
| Log2(MCP-1)  -0.366 | | 0.226 | 0.108 |  | | |
| Age >=65  0.937 | | 4.936 | 0.850 |  |  |  |
| Log2(MCP-1):Age >=65  -0.110 | | 0.535 | 0.838 |  |  |  |
|  | estimate | std.error | p.value | adj.r.squared | | |
| (Intercept) | 2.963 | 2.122 | 0.165 | 0.033 | | |
| Log2(MCP-1)  -0.208 | | 0.236 | 0.381 |  | | |
| Sex, Male  4.526 | | 4.333 | 0.299 |  |  |  |
| Log2(MCP-1):Sex, Male  -0.525 | | 0.472 | 0.268 |  |  |  |
| Log2(D28-D0) HAI response to vaccine strains by markers, adjusted for sex, age, vaccine dose | | | | | | |
| Response per strain | **Predictor** | **Adjusted** | **N** | **Estimate** | **Std.Error** | **P.Value** |
| Log2(HAI H1N1 Day 28/Day 0) | **CMV IgG Index** | **Sex, Age, Dose** | **320** | **0.099** | **0.042** | **0.02** |
| Log2(HAI H3N2 Day 28/Day 0) | **CMV IgG Index** | **Sex, Age, Dose** | **320** | **0.115** | **0.046** | **0.014** |
| Log2(HAI B/Vic Day 28/Day 0) | CMV IgG Index | Sex, Age, Dose | 320 | 0.072 | 0.04 | 0.074 |
| Log2(HAI H1N1 Day 28/Day 0) | Log2(MCP.1) | Sex, Age, Dose | 110 | -0.175 | 0.223 | 0.43 |
| Log2(HAI H3N2 Day 28/Day 0) | **Log2(MCP.1)** | **Sex, Age, Dose** | **110** | **-0.606** | **0.292** | **0.04** |
| Log2(HAI B/Vic Day 28/Day 0) | Log2(MCP.1) | Sex, Age, Dose | 110 | -0.248 | 0.218 | 0.26 |

**Supplementary Table 5. Distribution and comparisons of demographic variables and HAI responses among older adult recipients of SD vs HD influenza vaccines.** N-Miss, number of subjects with missing data; Q1, 25^th^ quartile; Q3, 75^th^ quartile; BMI, body mass index. Statistical comparisons between dose subgroups were evaluated using Wilcoxon rank sum tests.

| Demographic characteristics | | | |
| --- | --- | --- | --- |
|  | **Standard dose (SD)** | **High dose (HD)** | **p value** |
| Age |  |  | 0.818 |
| Median (Q1, Q3) | 69 (67, 72) | 70 (67, 72) |  |
| Range | 65 - 85 | 65 - 81 |  |
| Sex |  |  | 0.282 |
| F | 10 (47.6%) | 42 (60.9%) |  |
| M | 11 (52.4%) | 27 (39.1%) |  |
| Race |  |  | 0.283 |
| American Indian/Alaska Native | 0 (0.0%) | 1 (1.4%) |  |
| Asian | 0 (0.0%) | 1 (1.4%) |  |
| Black | 2 (9.5%) | 1 (1.4%) |  |
| White | 19 (90.5%) | 66 (95.7%) |  |
| BMI |  |  | 0.135 |
| N-Miss | 1 |  |  |
| Median (Q1, Q3) | 26.304 (25.497, 28.490) | 29.171 (24.913, 32.254) |  |
| Range | 19.708 – 39.619 | 19.082 – 52.14 |  |
| HAI responses | | | |
| maxRBA | 0.52 (-0.20, 2.67) | 0.23 (-0.20, 1.23) | 0.5 |
| Log2(HAI H1N1 Day 28/Day 0) | 1.00 (0.00, 2.00) | 1.00 (0.00, 2.00) | 0.6 |
| Log2(HAI H3N2 Day 28/Day 0) | 2.00 (0.00, 4.00) | 2.00 (1.00, 3.00) | 0.7 |
| Log2(HAI B/Vic Day 28/Day 0) | 1.00 (0.00, 3.00) | 1.00 (0.00, 2.00) | 0.2 |

**Supplementary Table 5. Effect of CMV, MCP-1, and IL-10 on vaccine-induced seroprotection.** Results of age- and sex-adjusted logistic regression models predicting seroprotection at D28 (HAI ≥ 1:40) in individuals who were seronegative to respective strains at baseline. N indicates the number of subjects seronegative at baseline with datapoints available for each marker of immunosenescence. SE, standard error.

| Influenza vaccine strain | Predictor/Marker |  | N | Estimate | SE | p value |
| --- | --- | --- | --- | --- | --- | --- |
| A/H1N1 | CMV IgG | Sex, Age, Dose | 237 | 0.02 | 0.015 | 0.194 |
|  | Log2(MCP.1) | Sex, Age, Dose | 65 | -0.161 | 0.125 | 0.201 |
|  | Log2(IL.10) | Sex, Age, Dose | 65 | -0.006 | 0.034 | 0.858 |
| A/H3N2 | **CMV IgG** | **Sex, Age, Dose** | **288** | **0.046** | **0.014** | **0.000961** |
|  | Log2(MCP.1) | Sex, Age, Dose | 102 | -0.08 | 0.088 | 0.36 |
|  | Log2(IL.10) | Sex, Age, Dose | 102 | -0.011 | 0.027 | 0.672 |
| B/Vic | CMV IgG | Sex, Age, Dose | 174 | 0.023 | 0.018 | 0.2 |
|  | Log2(MCP.1) | Sex, Age, Dose | 58 | 0.024 | 0.118 | 0.843 |
|  | Log2(IL.10) | Sex, Age, Dose | 58 | -0.058 | 0.041 | 0.16 |

**Supplementary Table 6. Effect of CMV serostatus on seroprotection induction (post-vaccination** HAI ≥ 1:40 in baseline HAI-seronegative (HAI < 1:40) individuals.

|  | CMV - (N=107) | CMV+ (N=132) | p value* |
| --- | --- | --- | --- |
| HAI H1N1 Seropositive at Day 28 |  |  | 0.2101 |
| No | 63 (58.9%) | 67 (50.8%) |  |
| Yes | 44 (41.1%) | 65 (49.2%) |  |
|  | Negative (N=138) | Positive (N=153) | p value |
| HAI H3N2 Seropositive at Day 28 |  |  | **0.0031** |
| No | 78 (56.5%) | 60 (39.2%) |  |
| Yes | 60 (43.5%) | 93 (60.8%) |  |
|  | Negative (N=70) | Positive (N=79) | p value |
| HAI B/Yam Seropositive at Day 28 |  |  | 0.4251 |
| No | 32 (45.7%) | 31 (39.2%) |  |
| Yes | 38 (54.3%) | 48 (60.8%) |  |
|  | Negative (N=88) | Positive (N=87) | p value |
| HAI B/Vic Seropositive at Day 28 |  |  | 0.9291 |
| No | 37 (42.0%) | 36 (41.4%) |  |
| Yes | 51 (58.0%) | 51 (58.6%) |  |
| *Pearson’s Chi-squared test |  |  |  |
